# Smarter, Shorter, Safer: Study protocol for a stepped-wedge cluster randomised controlled trial and process evaluation of a behavioural intervention bundle to reduce antibiotic use in residential aged care

**DOI:** 10.64898/2026.07.01.26357003

**Authors:** Magdalena Z. Raban, Rachel Urwin, Bayzidur Rahman, S. Sandun M. Silva, Ben R. Newell, Lyn-li Lim, Nasir Wabe, Ling Li, Gaston Arnolda, Sangita Neupane, Sarah Balmer, Travis Dunstan, Sonali Pinto, Verily Thomas, Johanna I. Westbrook

## Abstract

**Introduction:** The overuse and prolonged use of antibiotics in residential aged care (RAC) increases the risk of adverse effects and contributes to the global threat of antimicrobial resistance. Behavioural science-informed interventions have effectively reduced antibiotic prescribing in primary care but have rarely been implemented in RAC settings. We co-developed a behavioural science intervention bundle named “Smarter, Shorter, Safer” with RAC providers. We aim to assess the effectiveness of the intervention on rates of antibiotic use, measure effect persistence 6-months after the final intervention round, and investigate stakeholder experiences of the intervention.

**Methods and analysis:** We will conduct a stepped-wedge cluster randomised controlled trial with an embedded qualitative process evaluation. RAC homes (n=46) will be stratified into tertiles of baseline antibiotic use and randomly allocated to three intervention roll-out steps. Each RAC home will receive a bundled intervention consisting of social norm feedback, public commitment messaging and consumer information, staggered by step. Three rounds of the intervention will be delivered to each home at quarterly intervals. The primary outcome is antibiotic days of therapy per 1000 resident days (DOT/1000 days). Secondary outcomes are the percentage of antibiotic courses with a duration longer than guidelines and the percentage of residents on an antibiotic. Qualitative interviews will investigate staff and consumer experiences of the intervention bundle over time and by high, median and low baseline antibiotic use rates.

**Ethics and dissemination:** We obtained ethical approval from the Macquarie University Human Research Ethics Committee. Our findings will be disseminated through a range of forums including the publication of results in peer-reviewed journals and presentations at national and international conferences.

**Trial registration number:** Australian New Zealand Clinical Trials Registry ACTRN12625001132437 (https://anzctr.org.au/ACTRN12625001132437.aspx)

**Strengths and Limitations:**

- A low-cost “light touch” intervention bundle will support antimicrobial stewardship activities in RAC
- The social norm feedback component of the intervention is informed by best practice recommendations to optimise audit and feedback interventions and behavioural science.
- The longitudinal process evaluation comprising interviews will generate an understanding of intervention effect and whether experiences vary by homes’ rates of antibiotic use (low, median, high).
- The study will be conducted in 46 RAC homes in one country which may limit the generalisability of our findings to other contexts.

## Introduction

In residential aged care (RAC; also referred to as long term care, nursing homes and care homes), up to 70% of residents receive an antibiotic annually.(1, 2) However, there are high rates of inappropriate antibiotic use, defined as antibiotic use for infections where there is likely to be little or no benefit, and choice of an antibiotic, its dose or duration is not aligned with guidelines.(3, 4) Studies using surveillance criteria estimate that between 50-75% of antibiotic courses in RAC are inappropriate.(5-7) A recent analysis of antibiotic courses for urinary tract infections in RAC showed that 75% of courses were of longer duration than recommended by guidelines.(8) Another analysis showed that only 28% of antibiotic courses prescribed for suspected urinary tract infections were with guideline recommended antibiotics.(7)

Inappropriate antibiotic use increases the risk of adverse effects. Older adults living in RAC are at higher risk of adverse effects due to polypharmacy, multimorbidity and age-related physiological changes.(9, 10) Importantly, overuse of antibiotics leads to increases in antimicrobial resistance, recognised as a leading threat to global human health by the World Health Organisation.(11) Thus, addressing the overuse of antibiotics is a key priority in RAC globally. In Australia, the Aged Care Quality Standards recognise the pressing need for antimicrobial stewardship in RAC and require RAC homes to ensure judicious antibiotic use.(12)

Behavioural science informed interventions have successfully reduced antibiotic prescribing in primary care.(13, 14) Strategies employed have included social norm feedback, i.e. providing participants with feedback on their antibiotic prescribing practices relative to their peers and public commitment.(15, 16) Social norm feedback interventions, also termed audit and feedback, in primary care have taken advantage of large administrative datasets on antibiotic prescribing and dispensing to provide feedback to general practitioners.(17, 18)

In RAC, interventions to reduce antibiotic use have primarily involved educational strategies and guideline implementation.(19, 20) No studies have employed other behavioural science informed interventions in RAC.(19-21) A recent systematic review of 12 studies of antibiotic use interventions in RAC reported that there was low quality evidence that interventions led to reductions in antibiotic prescriptions.(19) Of the 12 studies, five studies included an audit and feedback component; however, they were conducted over 10 years ago or contained minimal description of how the feedback was designed and delivered.(22-26)

Two large meta-analyses of audit and feedback interventions in healthcare, published in 2012 and 2025, identified characteristics of feedback to optimise their effectiveness in driving changes in practice.(27, 28) Furthermore, feedback interventions can draw on evidence from psychology and behavioural economics to inform the design of effective feedback.(29, 30) Despite this emerging evidence-base, key questions remain about the effectiveness of social norm feedback interventions in RAC. For example, it is unknown whether providing social norm feedback at the RAC home level, rather than to prescribers, will result in reductions in use, despite RAC staff having an established role in decisions to prescribe antibiotics.(31, 32)

In 2022 we established a National Aged Care Medication Roundtable with RAC providers to support data driven improvements in medication management.(33) As part of this work, we aimed to co-develop and test behavioural interventions to support improved antibiotic use.(34) Our co-development work with our aged care provider partners indicated that a bundled intervention consisting of social norm feedback, public commitment and consumer information would be acceptable and feasible.

### Objectives

Our aims are to: 1) Assess the effectiveness of an intervention bundle with repeated social norm feedback (3 rounds), public commitment and consumer information on reducing antibiotic use in RAC; 2) Assess intervention effect persistence 6-months after the final round of social norm feedback; and 3) Understand stakeholder experiences of the intervention bundle, whether these vary by baseline performance (low, average, high antibiotic use) and whether they change over time.

## Methods and Analysis

This protocol is reported according to the Standard Protocol Items for Randomized Trials (SPIRIT) statement (checklist in Supplementary file 1).(35)

### Patient and public involvement

The materials included in the intervention bundle were co-designed with members of the National Aged Care Medication Roundtable through stakeholder intervention planning meetings. We consulted extensively with RAC provider representatives regarding the effective delivery of the intervention bundle, and the feasibility and acceptability of the trial design. Older adults living in RAC were not involved in the design of this study, but will participate in the evaluation of its effects.

### Study design

This is a stepped-wedge cluster randomised controlled trial (SWCRCT) with a parallel qualitative process evaluation and a 6-month follow-up measurement period to assess intervention effect persistence (Figure 1).

**Figure 1.**
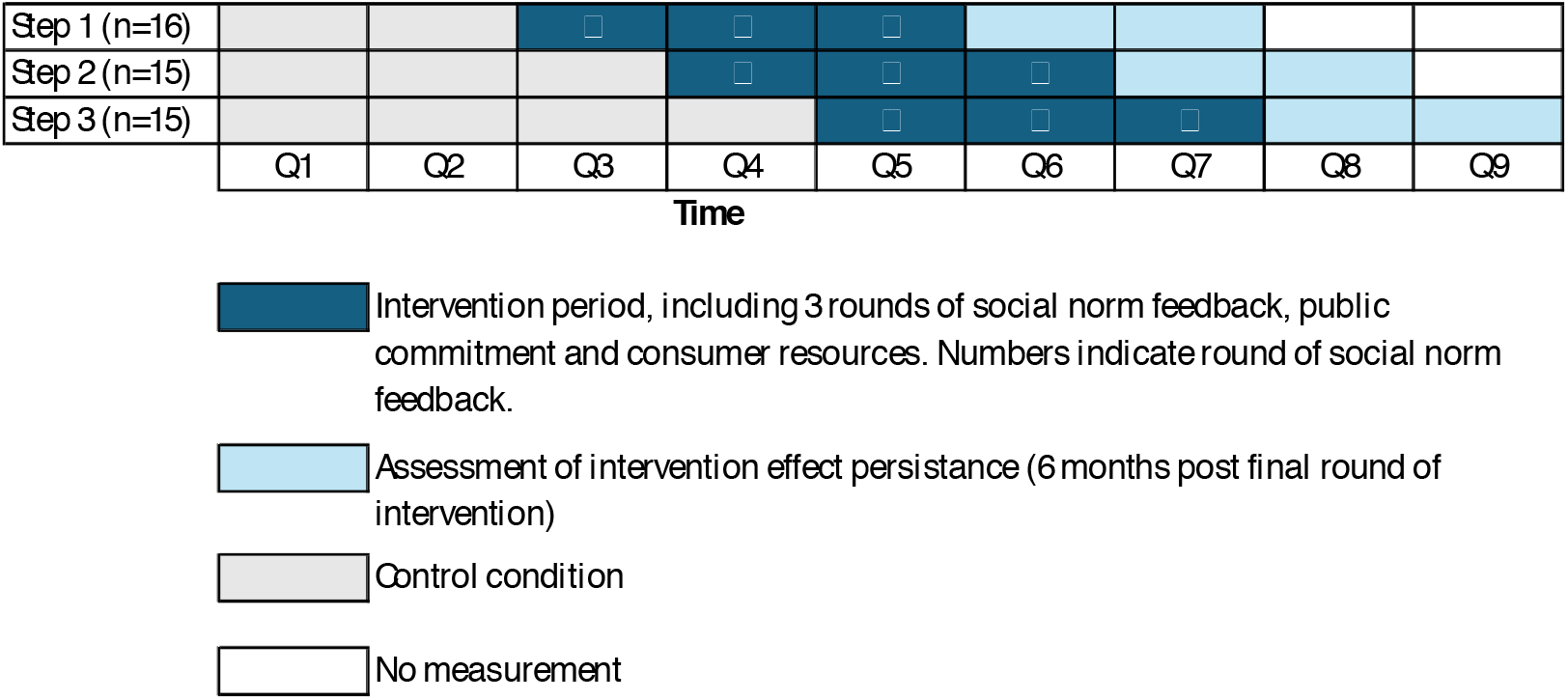
Stepped-wedge cluster randomised controlled trial design. Q is quarter. N represents the number of residential aged care homes.

During the 15-month trial period, the intervention bundle will be rolled out sequentially to 46 RAC homes (clusters) in three steps, with each home receiving three rounds of the intervention (Figure 1). There will be a pre-intervention period of a minimum of 6 months (9 months for Step 2 homes and 12 months for Step 3 homes) to establish outcome trends prior to the intervention.

Semi-structured interviews will be conducted in a sub-sample of intervention RAC homes at 3-months and 9-months following the first round of feedback to assess stakeholder experiences and perceived effects of the intervention bundle and how these change over time.

### Trial setting and eligibility criteria

This study will be conducted in partnership with three RAC service providers in New South Wales, Australia, who are members of the National Aged Care Medication Roundtable (22, 18 and 6 homes managed by provider A, B and C, respectively).(33) The RAC homes are of varying sizes, ranging from 48 to 172 beds and varying baseline use of antibiotics ranging from 41.7 to 145.3 per 1000 resident days (as of quarter 2, 2025).

RAC homes provide care to older adults who are no longer able to manage living in their own home. Medication prescribing for older adults residing in RAC is undertaken by general practitioners (GPs) and nurse practitioners. Medicines are dispensed by a supplying pharmacy and administered in the RAC home by aged care staff or self-administered by residents. GPs and supplying pharmacies are located off-site in the community and when a resident is unwell, the GP will be contacted by the RAC nurse or other staff. Regular Medication Advisory Committee meetings are held with RAC staff, GPs and pharmacists to discuss medication management issues.

#### Eligibility

All RAC homes will be eligible to participate in the SWCRCT.

#### Process evaluation eligibility

We will conduct interviews with staff from RAC homes at 3 and 9 months after the first round of the intervention. To identify interviewees, we will purposively sample participants from RAC homes based on the homes’ baseline rates of antibiotic use, as measured by antibiotic days of therapy per 1000 resident days (DOT/1000 days). RAC staff, residents and their families and carers from RAC homes with low (lowest quartile), average (close to median) and high antibiotic (highest quartile) use will be invited to participate.

### Intervention and comparator

The intervention will include: 1) social norm feedback about antibiotic use in the RAC home; 2) public commitment; 3) consumer information. During the control period, care will be delivered as usual. The intervention bundle was co-developed with the aged care providers through a consultation process and workshops. The bundle of interventions will be delivered to all sites in a staggered manner for a total of three rounds at quarterly intervals. The campaign has been named “Smarter, Shorter, Safer” with the messaging consistent across all intervention components. Figure 2 shows the campaign logo.

**Figure 2.**
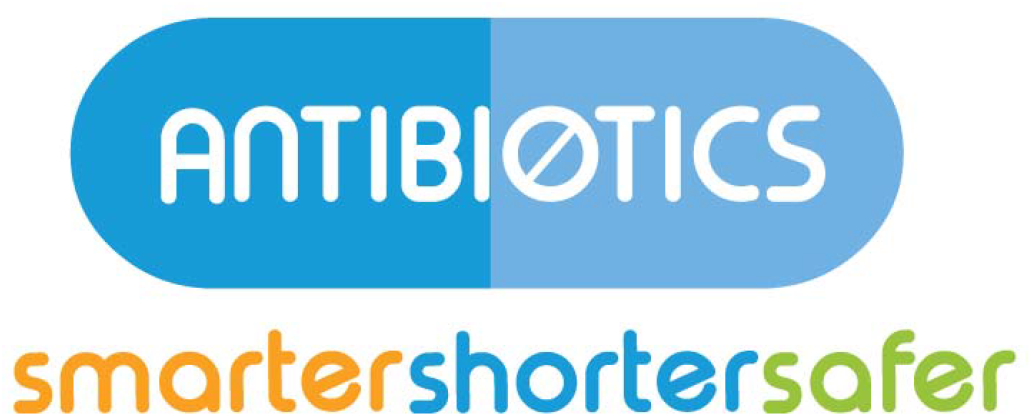
Logo used across all intervention materials

All intervention components will be delivered via email and letter, addressed to the RAC home’s manager. Thus, intervention homes will receive electronic and hard copies of the materials. One week prior to the first intervention round, an email will be sent to the RAC managers to inform them that they will receive the intervention. Additionally, half-way through the first and second social norm feedback quarter, an email will be sent to RAC managers in the intervention group to remind them that another round of feedback will be delivered in a few weeks. A description of the intervention components is provided below.

#### Social norm feedback

Social norm feedback will be delivered in the form of a letter and email addressed to the RAC home’s residential manager on a quarterly basis. The content of the letter was informed by guidelines for the design of audit and feedback interventions(36, 37) (detailed considerations shown in Supplementary file 2) and evidence on social norm feedback from the behavioural economics literature.(34) The key elements to support the effectiveness of social norm feedback will be: i) a performance target; ii) actionable advice to improve performance; iii) an injunctive norm; iv) a respected signatory; v) multiple rounds of feedback.

#### Data presented and performance target

Electronic medication administration data from each RAC home will be used to generate the social norm feedback. The data contain information on each medication dose administered to each resident and will be used to generate a customised letter for each RAC home about their antibiotic use. The main message contained in the letter will be a graphical presentation of the monthly rate of systemic antibiotic use (DOT/1000 days) for the home for the preceding three months, compared to the target performance. The target performance will be the mean of the lowest decile of DOT/1000 days of RAC homes receiving the intervention. The target performance will be calculated for every quarter, with the homes included in the calculation varying based on the number of rounds of feedback they have received. The target performance will be drawn from homes that have received the same number of rounds of the intervention. All estimates of DOTs/1000 days will be risk adjusted to account for differences in resident age, gender and health conditions using a regression-based indirect standardisation method.(38)

#### Actionable advice

The customised letter will contain two actions that can be taken to reduce the DOT/1000 days. Firstly, the feedback will contain the message that ‘Shorter is better’.(39) That is, when antibiotics are used, they should be used for a duration of 5 days for most infections.(40, 41) The messaging is based on an intervention targeting primary care doctors’ antibiotic prescribing for older adults (aged ≥ 65 years) in Canada.(39) The duration of 5 days was informed by the Australian Therapeutic Guidelines on Antibiotics.(41) The percentage of antibiotic courses in the RAC home during the last quarter that were longer than guidelines, along with the guideline recommendations for duration of treatment for common infections will be presented.(41)

Secondly, there will be a message for ‘smarter’ and ‘safer’ use, encouraging reduced use of antibiotics, with a reminder that antibiotics may cause adverse effects and are ineffective against key infections (e.g. viral infections). Letters will also suggest how the information can be communicated to key stakeholders e.g. through presentation at medication advisory committee meetings.

#### Injunctive norm

We will provide clear messaging about whether the home’s performance is perceived positively or negatively. There will be three messages based on the home’s level of antibiotic use. The comparison antibiotic use rates, i.e. the target performance, will be described as the ‘best performing’ homes. RAC homes with rates of antibiotic use (DOT/1000 days) within the lowest decile will be provided with a message stating ‘you are a best performer’ to support well performing homes to maintain their performance. RAC homes with DOT/1000 days in the lowest quartile (bottom 25%), but which are not currently matching the best performers, will be provided a message stating ‘you are close to being a best performer’. The remainder of homes will have the message that their antibiotic use is ‘higher’ than best performing homes. All feedback will include a message reinforcing that ‘lower is better’.

#### Respected signatory

The social norm feedback letter signatories will be unique to each aged care provider and will be clinical governance leaders from within each organisation. This will increase the credibility of the feedback through local leadership support.

#### Multiple rounds of feedback

Each RAC home will receive a total of 3 rounds of feedback.

#### Public commitment

Poster-sized commitment messages will be provided to the intervention homes to be displayed in staff areas and common spaces within each RAC home, once the intervention has been implemented (i.e. the delivery of social norm feedback has begun in the home). Posters will feature aged care staff photographs and signatures and will emphasise the organisation’s commitment to best practice use of antibiotics, including the use of antibiotics only when necessary and following treatment guidelines on the length of time antibiotics are used.

#### Consumer information

Information will be displayed and distributed to consumers (residents and their families) via digital screens and notice boards in common areas, newsletters, and leaflets. Simple messages will be promoted that are consistent with key elements of the social norm feedback and public commitment, including the organisational commitment to use antibiotics only when necessary and following treatment guidelines on the length of time they are used. A link to additional resources (42) will be accessible via a QR code included on consumer-focussed materials.

### Outcomes

The primary outcome will be the change in the mean antibiotic days of therapy per 1000 resident days (DOT/1000 days) from baseline across all RAC homes. Secondary outcomes will be the change in the percentage of antibiotic courses with a duration longer than guidelines and the percentage of residents on an antibiotic.

#### Process evaluation

The qualitative interviews will provide RAC staff and consumer perspectives on, and action taken as a result of, the intervention bundle, how these change over time and how they vary between stakeholders of homes with low, average and high antibiotic use.

### Sample size

Based on pilot data, we assumed an antibiotic use rate of 76 DOT/1000 days in the control condition. With an average cluster size of 104 (SD: 33) residents and an intraclass correlation coefficient (ICC) of 0.01, a 1.8% minimum detectable difference post-intervention for the primary outcome with a power of 80% and 5% significance level.(43) Given the absence of prior ICC estimates for this setting, a conservative value of 0.01 was assumed, consistent with empirical evidence that ICCs in clinical cluster randomised trials are generally small.(44)

For the secondary outcome of proportion of courses longer than guidelines, we will be able to detect a 1.48% change with a power of 80% and at a 5% significance level.

Uncontrolled before and after studies of interventions in RAC reported declines in antibiotic DOT/1000 days ranging from 20% to 35%.(45-47) Social norm feedback interventions are expected to produce a mean increase in desired practice of 6.2%.(27) Thus, the study will have adequate statistical power with 46 participating RAC homes.

### Allocation

An independent statistician who will not be involved in implementation, data collection, or analysis will perform the randomisation. Randomisation will occur once at the start of the trial.

Participating RAC homes (n=46) will first be stratified into tertiles according to risk adjusted baseline DOT/1000 resident days: low use (lower tertile: 41.7-58 DOT/1000 resident days), average use (middle range: 58.1-89.8 DOT/1000 resident days), and high use (upper tertile: 89.9-145.3 DOT/1000 resident days). Then from each tertile, RAC homes will be equally allocated to one of three steps, as shown in Figure 1.

RAC homes within each tertile will be equally allocated across the intervention time steps. This will be done using a fixed seed number to enhance the reproducibility and transparency. The random allocation of 15-16 homes to one of the sequential steps will determine the timing at which each group of RAC homes transitions from the control to intervention condition.

#### Allocation concealment

The randomisation sequence will be generated and maintained by an independent researcher not involved in implementation (or recruitment). This will ensure allocation concealment prior to trial commencement and minimise selection bias. Due to the nature of the SWCRT design, allocation cannot be concealed after rollout begins; however, intervention status will be coded to maintain blinding of the statistician conducting the analyses.

### Blinding

RAC homes will not be blinded to their allocation, i.e. they will be aware they have received the intervention; however, they will not be aware of the order in which they will receive the intervention. RAC home staff will be informed that they will receive the intervention one week before the implementation of the intervention via email.

### Data collection and measurement

#### SWCRCT

Electronic health record data will be extracted on a quarterly basis from aged care providers’ information systems, including medication administration records (e.g., the medicines each resident receives daily) and resident profile data which comprises demographic information (e.g., age, sex), and health conditions (e.g., dementia, Parkinson’s disease). We will apply the World Health Organization’s Anatomical Therapeutic Classification (ATC) system to identify systemic (oral and injectable) antibiotics that have been prescribed and administered to residents.

##### DOT/1000 days definition

Measurement of antibiotic DOT/1000 resident days will follow established methods.(48, 49) Briefly, DOT will be calculated using the medication administration data by counting the number of days of administration of each antibiotic. If two different antibiotics are administered on one day to one resident, these will be counted separately. The antibiotics included in the DOT will be all systemic antibiotics and will be identified using WHO Anatomical Therapeutic Chemical classification codes, specifically J01 (except J01XX05). The number of days each resident is present in a RAC home will be extracted from the resident profile dataset.

##### Proportion of courses longer than guidelines

The proportion of antibiotic courses longer than guidelines will be calculated based on key prescribing guidelines for common infections as provided by the Therapeutic Guidelines: Antibiotics.(41) This includes antibiotic use for treatment of infections and prophylaxis of urinary tract infections. Table 2 shows the list of antibiotics and treatment lengths identified as longer than recommended for common infections in RAC such as cystitis, community-acquired pneumonia and mild cellulitis.

**Table 2.**
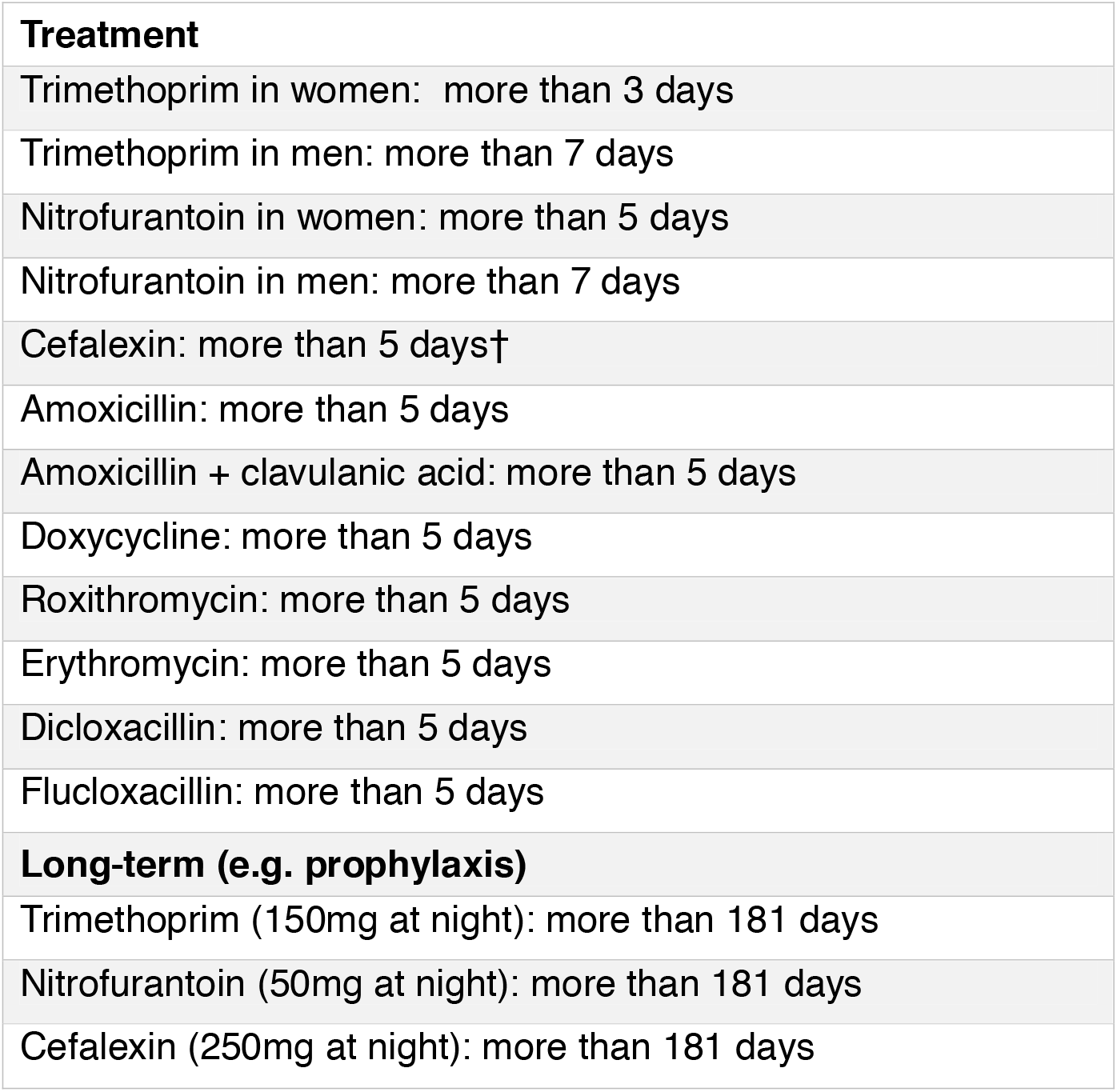
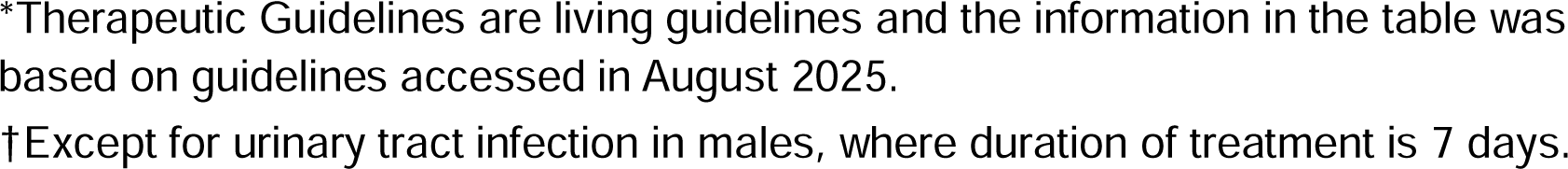
Antibiotic course lengths classified as longer than recommended*(41)

|  |
| --- |
| **Treatment** |
| Trimethoprim in women: more than 3 days |
| Trimethoprim in men: more than 7 days |
| Nitrofurantoin in women: more than 5 days |
| Nitrofurantoin in men: more than 7 days |
| Cefalexin: more than 5 days† |
| Amoxicillin: more than 5 days |
| Amoxicillin + clavulanic acid: more than 5 days |
| Doxycycline: more than 5 days |
| Roxithromycin: more than 5 days |
| Erythromycin: more than 5 days |
| Dicloxacillin: more than 5 days |
| Flucloxacillin: more than 5 days |
| **Long-term (e.g. prophylaxis)** |
| Trimethoprim (150mg at night): more than 181 days |
| Nitrofurantoin (50mg at night): more than 181 days |
| Cefalexin (250mg at night): more than 181 days |

\*Therapeutic Guidelines are living guidelines and the information in the table was based on guidelines accessed in August 2025.
†Except for urinary tract infection in males, where duration of treatment is 7 days.

#### Process evaluation

We will recruit RAC staff with clinical, governance or managerial roles (e.g. RNs, residential managers, care managers), as well as consumers (residents and carers) via email and during site visits. After providing informed consent, participants will complete 30-minute semi-structured interviews which will be audio-recorded. The interview questions will relate to each component of the intervention bundle: audit and feedback, public commitment and consumer poster, and are provided in Supplementary file 3. The questions related to audit and feedback questions materials were informed by a process evaluation of two audit and feedback trials in Canada.(50)

## Statistical methods and data analysis

### SWCRCT

The intention-to-treat analysis approach will be adopted to examine the effectiveness of the intervention on the outcome variables, and the effect measures will be reported as rate ratio/risk ratio (RR) or incidence rate ratio (IRR). Multilevel mixed effects generalised linear models will be applied with robust standard error to estimate the main effect. Clusters (RAC homes) will be included as a random factor and calendar time (of rolling out the intervention) as a fixed factor along with a time-intervention interaction in the model. Other potential confounders at patient level (e.g., age, sex, co-morbidities, etc) as well as at cluster level (e.g., size of the RAC home, geographical location, etc) will be adjusted for in the model. If a significant and clinically important time-intervention interaction is found, we will estimate the time-specific effect of the intervention on the outcomes. Under this framework, a multilevel negative binomial model will be adopted for our primary outcome (days of therapy of antibiotic use/ 1000 resident days) to estimate IRR, and multilevel log-binomial/Poisson models for the secondary outcome (proportion of antibiotic courses with a duration longer than guidelines) to estimate risk ratio.

To assess the persistence of the intervention effect, we will examine the trend of antibiotic use for 6 months after the intervention period is over. In this analysis, we will model monthly antibiotic use in a cluster (RAC homes) using multivariable generalised linear model to adjust for potential confounders. For the purposes of comparison, data from 6 months prior to the end of intervention will be included. Trend lines before and after the intervention end point will be compared as the main effect. If a non-linear trend is observed a restricted cubic spline approach will be adopted to model the time and generate the adjusted trend line. To model the rate of antibiotic use (DOT/1000 resident days), a multilevel Poisson/negative binomial model will be applied. For proportion of antibiotic use (binary outcome), a log-binomial/Poisson model will be applied under the same framework to estimate IRR and RR, respectively. Adjustment will be made for all the relevant covariates, as mentioned in the previous paragraph, in all the models.

### Process evaluation

Qualitative data of consumer, care staff, home manager perspectives of each intervention component will be analysed using inductive content analysis. Two researchers will independently perform open coding of three to five transcripts and then meet to develop a preliminary coding framework through discussion. To develop the preliminary coding framework, codes will be sorted into domains, themes and subthemes. This framework will then be applied to analysis of a sample of subsequent transcripts and refined to maximise homogeneity in a final coding framework. The final coding framework will be applied to all transcripts and lead to the development of an analytic narrative of interview results. A third reviewer will resolve any disagreements at any point to reach consensus. Data will be compiled to determine stakeholder experiences of the intervention bundle overall, relative to the RAC homes antibiotic use level (low, median, high) and how they change over time.

### Data monitoring

A systematic review of antibiotic stewardship interventions in RAC found interventions were safe with no impact on hospitalisation and mortality.(19) Furthermore, the intervention is delivered to RAC homes, rather than individual residents, with the aim of supporting antimicrobial stewardship activities. Usual antimicrobial stewardship activities in the RAC homes will continue. Thus, as the trial is low-risk, trial monitoring will be conducted via the Project Steering Committee and Roundtable. The Project Steering Committee consists of the project team, Chief Investigators, and partner representatives (including from RAC providers), and meets quarterly. The Roundtable meets quarterly and consists of the project team and RAC partner representatives, with some meetings also attended by RAC home staff. At these meetings, updates will be provided by the project team regarding trial progress and RAC partners will provide updates and feedback from their RAC homes on the intervention.

## Ethics and dissemination

Ethics for this study was granted by the Macquarie University Human Research Ethics Committee (Ref: 520221126736854). The approval included a waiver for individual consent in accordance with the National Statement on Ethical Conduct in Human Research,(51) including the intervention being delivered to RAC homes, rather than individual residents, and relying on retrospective de-identified electronic health record data for evaluation.

Trial results will be presented to aged care providers and project partners, and disseminated in peer-reviewed journal articles and conference presentations.

## Data Availability

This is a trial protocol and there are no data available.

## Authors’ contributions

MZR, JIW and BN conceptualised the study. MZR, JIW, RU, SMS, BN, NW, LLL, LL, SN, SP, TD, VT, SB designed the intervention with input and in collaboration with aged care providers. All authors contributed to the trial design. BR led the statistical analysis plan, supported by GA. MZR led the manuscript drafting. All authors reviewed and approved the final version of the manuscript.

## Funding statement

This study is supported by a National Health and Medical Research Council Partnership Projects Grant (c). JIW is supported by an NHMRC Investigator Leadership Fellowship (APP2032961).

## Competing interest statement

The authors have no competing interests.

## Notes

### Competing Interest Statement

The authors have declared no competing interest.

### Clinical Trial

ACTRN12625001132437

### Author Declarations

Ethics for this study was granted by the Macquarie University Human Research Ethics Committee (Ref: 520221126736854).

## References

1. Raban MZ, Gates PJ, Gasparini C, Westbrook JI. Temporal and regional trends of antibiotic use in long-term aged care facilities across 39 countries, 1985-2019: Systematic review and meta-analysis. PLoS One. 2021;16(8):e0256501.

2. Raban MZ, Lind KE, Day RO, Gray L, Georgiou A, Westbrook JI. Trends, determinants and differences in antibiotic use in 68 residential aged care homes in Australia, 2014–2017: a longitudinal analysis of electronic health record data. BMC Health Services Research. 2020;20(1):883.

3. Otaigbe, II, Elikwu CJ. Drivers of inappropriate antibiotic use in low- and middle-income countries. JAC Antimicrob Resist. 2023;5(3):dlad062.

4. Laka M, Milazzo A, Merlin T. Inappropriate antibiotic prescribing: understanding clinicians’ perceptions to enable changes in prescribing practices. Australian Health Review. 2022;46(1):21–7.

5. Gyawali R, Gamboa S, Rolfe K, Westbrook JI, Raban MZ. Consumer perspectives on antibiotic use in residential aged care: A mixed-methods systematic review. American Journal of Infection Control. 2024.

6. Falconer N, Paterson DL, Peel N, Welch A, Freeman C, Burkett E, et al. A multimodal intervention to optimise antimicrobial use in residential aged care facilities (ENGAGEMENT): protocol for a stepped-wedge cluster randomised trial. Trials. 2022;23(1):427.

7. Hansen MB, Lykkegaard J, Hansen MP, Llor C, Sangenis AG, Touboul-Lundgren P, et al. Appropriateness of antibiotic use in nursing homes for suspected urinary tract infections: comparison across five European countries. Eur Geriatr Med. 2025;16(4):1453–64.

8. Raban M, Gyawali R, Nguyen A, Lim LL, Stewart K, Westbrook J. Designing a Clinical Decision Support Prototype for Urinary Tract Infection Treatment in Nursing Homes: Experiences from a Research Partnership. Stud Health Technol Inform. 2025;327:158–62.

9. Sluggett JK, Harrison SL, Ritchie LA, Clough AJ, Rigby D, Caughey GE, et al. High-Risk Medication Use in Older Residents of Long-Term Care Facilities: Prevalence, Harms, and Strategies to Mitigate Risks and Enhance Use. The Senior Care Pharmacist. 2020;35(10):419–33.

10. Fried TR, O’Leary J, Towle V, Goldstein MK, Trentalange M, Martin DK. Health outcomes associated with polypharmacy in community-dwelling older adults: a systematic review. J Am Geriatr Soc. 2014;62(12):2261–72.

11. World Health Organization. Antimicrobial resistance 2023 [Available from: https://www.who.int/news-room/fact-sheets/detail/antimicrobial-resistance#:~:text=Antimicrobial%20Resistance%20(AMR)%20occurs%20when,systems%20and%20national%20economies%20overall.

12. Australian Commission on Safety and Quality in Health Care. Antimicrobial Stewardship in Australian Health Care. 2021.

13. Tonkin-Crine S, Walker AS, Butler CC. Contribution of behavioural science to antibiotic stewardship. BMJ. 2015;350:h3413.

14. Maher A, Roche K, Morrissey E, Murphy A, Sheaf G, Ryan C, et al. Behaviour change interventions addressing patient antibiotic treatment-seeking behaviour for respiratory tract infections in primary and community care settings: a scoping review. BMJ Open. 2025;15(8):e101694.

15. Bocquier A, Essilini A, Pereira O, Welter A, Pulcini C, Thilly N. Impact of a public commitment charter, a non-prescription pad and an antibiotic information leaflet to improve antibiotic prescription among general practitioners: A randomised controlled study. Journal of Infection and Public Health. 2024;17(2):217–25.

16. Raban MZ, Gonzalez G, Nguyen AD, Newell BR, Li L, Seaman KL, et al. Nudge interventions to reduce unnecessary antibiotic prescribing in primary care: a systematic review. BMJ Open. 2023;13(1):e062688.

17. Hallsworth M, Chadborn T, Sallis A, Sanders M, Berry D, Greaves F, et al. Provision of social norm feedback to high prescribers of antibiotics in general practice: a pragmatic national randomised controlled trial. The Lancet. 2016;387(10029):1743–52.

18. Ratajczak M, Gold N, Hailstone S, Chadborn T. The effectiveness of repeating a social norm feedback intervention to high prescribers of antibiotics in general practice: a national regression discontinuity design. J Antimicrob Chemother. 2019;74(12):3603–10.

19. Crespo-Rivas JC, Guisado-Gil AB, Peñalva G, Rodríguez-Villodres Á, Martín-Gandul C, Pachón-Ibáñez ME, et al. Are antimicrobial stewardship interventions effective and safe in long-term care facilities? A systematic review and meta-analysis. Clin Microbiol Infect. 2021;27(10):1431–8.

20. Raban MZ, Gasparini C, Li L, Baysari MT, Westbrook JI. Effectiveness of interventions targeting antibiotic use in long-term aged care facilities: a systematic review and meta-analysis. BMJ Open. 2020;10(1):e028494.

21. Vicentini C, Libero G, Cugudda E, Gardois P, Zotti CM, Bert F. Barriers to the implementation of antimicrobial stewardship programmes in long-term care facilities: a scoping review. J Antimicrob Chemother. 2024;79(8):1748–61.

22. Nace DA, Hanlon JT, Crnich CJ, Drinka PJ, Schweon SJ, Anderson G, et al. A Multifaceted Antimicrobial Stewardship Program for the Treatment of Uncomplicated Cystitis in Nursing Home Residents. JAMA Intern Med. 2020;180(7):944–51.

23. Pasay DK, Guirguis MS, Shkrobot RC, Slobodan JP, Wagg AS, Sadowski CA, et al. Antimicrobial stewardship in rural nursing homes: Impact of interprofessional education and clinical decision tool implementation on urinary tract infection treatment in a cluster randomized trial. Infect Control Hosp Epidemiol. 2019;40(4):432–7.

24. Sloane PD, Zimmerman S, Ward K, Kistler CE, Paone D, Weber DJ, et al. A 2-Year Pragmatic Trial of Antibiotic Stewardship in 27 Community Nursing Homes. J Am Geriatr Soc. 2020;68(1):46–54.

25. Monette J, Miller MA, Monette M, Laurier C, Boivin J-F, Sourial N, et al. Effect of an Educational Intervention on Optimizing Antibiotic Prescribing in Long-Term Care Facilities. Journal of the American Geriatrics Society. 2007;55(8):1231–5.

26. Zimmerman S, Sloane PD, Bertrand R, Olsho LEW, Beeber A, Kistler C, et al. Successfully Reducing Antibiotic Prescribing in Nursing Homes. Journal of the American Geriatrics Society. 2014;62(5):907–12.

27. Ivers N, Yogasingam S, Lacroix M, Brown KA, Antony J, Soobiah C, et al. Audit and feedback: effects on professional practice. Cochrane Database of Systematic Reviews. 2025(3).

28. Ivers N, Jamtvedt G, Flottorp S, Young JM, Odgaard-Jensen J, French SD, et al. Audit and feedback: effects on professional practice and healthcare outcomes. Cochrane Database Syst Rev. 2012;2012(6):Cd000259.

29. Thaler R, Sunstein C. NUDGE: Improving Decisions About Health, Wealth, and Happiness2009.

30. Schultz PW, Nolan JM, Cialdini RB, Goldstein NJ, Griskevicius V. The constructive, destructive, and reconstructive power of social norms. Psychol Sci. 2007;18(5):429–34.

31. Lim CJ, Kwong MW, Stuart RL, Buising KL, Friedman ND, Bennett NJ, et al. Antibiotic prescribing practice in residential aged care facilities--health care providers’ perspectives. Med J Aust. 2014;201(2):98–102.

32. Singh S, Degeling C, Fernandez D, Montgomery A, Caputi P, Deane FP. How do aged-care staff feel about antimicrobial stewardship? A systematic review of staff attitudes in long-term residential aged-care. Antimicrob Resist Infect Control. 2022;11(1):92.

33. Westbrook JI, Seaman K, Wabe N, Raban MZ, Urwin R, Badgery-Parker T, et al. Designing an Informatics Infrastructure for a National Aged Care Medication Roundtable. Stud Health Technol Inform. 2024;310:404–8.

34. Beatty A MR, Buttenheim A,. The Behavioral Economics Toolkit: Policy Levers and Intervention Strategies. Behavioral Economics: Policy Impact and Future Directions: National Academies Press; 2023.

35. Hróbjartsson A, Boutron I, Hopewell S, Moher D, Schulz KF, Collins GS, et al. SPIRIT 2025 explanation and elaboration: updated guideline for protocols of randomised trials. British Medical Journal. 2025;389:e081660.

36. Jamtvedt G, Flottorp, S, Ivers N. Audit and Feedback as a Quality Strategy. Improving healthcare quality in Europe: Characteristics, effectiveness and implementation of different strategies. Health Policy Series. 53 ed. Copenhagen (Denmark): European Observatory on Health Systems and Policies; 2019.

37. Langford BJ, Schwartz KL. Audit and feedback to improve antibiotic prescribing in primary care—the time is now. BMJ Quality & Safety. 2025;34(5):282–4.

38. Wattier RL, Thurm CW, Parker SK, Banerjee R, Hersh AL. Indirect Standardization as a Case Mix Adjustment Method to Improve Comparison of Children’s Hospitals’ Antimicrobial Use. Clin Infect Dis. 2021;73(5):925–32.

39. Schwartz KL, Shuldiner J, Langford BJ, Brown KA, Schultz SE, Leung V, et al. Mailed feedback to primary care physicians on antibiotic prescribing for patients aged 65 years and older: pragmatic, factorial randomised controlled trial. Bmj. 2024;385:e079329.

40. Llewelyn MJ, Fitzpatrick JM, Darwin E, SarahTonkin-Crine Gorton C, Paul J, et al. The antibiotic course has had its day. BMJ. 2017;358:j3418.

41. Therapeutic Guidelines Limited. Antibiotic prescribing in primary care: Therapeutic Guidelines summary table 2025. May 2025.

42. Aged Care Quality and Safety Commission. Do you need antibiotics? 2024 [Available from: https://www.agedcarequality.gov.au/resource-library/do-you-need-antibiotics.

43. Hemming K, Haines TP, Chilton PJ, Girling AJ, Lilford RJ. The stepped wedge cluster randomised trial: rationale, design, analysis, and reporting. BMJ : British Medical Journal. 2015;350:h391.

44. Heagerty P, DeLong E, for the NIH Health Care Systems Research Collaboratory Biostatistics and Study Design Core. Analysis Plan: Intraclass Correlation. Rethinking Clinical Trials: A Living Textbook of Pragmatic Clinical Trials. Bethseda: NIH Health Care Systems Research Collaboratory; 2024.

45. van Buul LW, van der Steen JT, Achterberg WP, Schellevis FG, Essink RT, de Greeff SC, et al. Effect of tailored antibiotic stewardship programmes on the appropriateness of antibiotic prescribing in nursing homes. J Antimicrob Chemother. 2015;70(7):2153–62.

46. Fleet E, Gopal Rao G, Patel B, Cookson B, Charlett A, Bowman C, et al. Impact of implementation of a novel antimicrobial stewardship tool on antibiotic use in nursing homes: a prospective cluster randomized control pilot study. J Antimicrob Chemother. 2014;69(8):2265–73.

47. Sloane PD, Zimmerman S, Reed D, Beeber AS, Chisholm L, Kistler C, et al. Antibiotic prescribing in 4 assisted-living communities: incidence and potential for improvement. Infect Control Hosp Epidemiol. 2014;35 Suppl 3:S62–8.

48. Barlam TF, Cosgrove SE, Abbo LM, MacDougall C, Schuetz AN, Septimus EJ, et al. Implementing an Antibiotic Stewardship Program: Guidelines by the Infectious Diseases Society of America and the Society for Healthcare Epidemiology of America. Clin Infect Dis. 2016;62(10):e51–77.

49. Polk RE, Fox C, Mahoney A, Letcavage J, MacDougall C. Measurement of adult antibacterial drug use in 130 US hospitals: comparison of defined daily dose and days of therapy. Clin Infect Dis. 2007;44(5):664–70.

50. Shuldiner J, Lacroix M, Saragosa M, Reis C, Schwartz KL, Gushue S, et al. Process evaluation of two large randomized controlled trials to understand factors influencing family physicians’ use of antibiotic audit and feedback reports. Implementation Science. 2024;19(1):65.

51. National Health and Medical Research Council, Australian Research Council, Universities Australia. National Statement on Ethical Conduct in Human Research. Canberra: National Health and Medical Research Council; 2025.

