## Supplementary file for "Smarter, Shorter, Safer: Study protocol for a stepped-wedge cluster randomised controlled trial and process evaluation of a behavioural intervention bundle to reduce antibiotic use in residential aged care"

### Supplementary file 1: Standard Protocol Items for Randomized Trials checklist

| Section / Topic | No | SPIRIT 2025 checklist item description | Reported on page no. |
|---|---|---|---|
| **Administrative information** | | | |
| Title and structured summary | 1a | Title stating the trial design, population, and interventions, with identification as a protocol | 1 |
| | 1b | Structured summary of trial design and methods, including items from the World Health Organization Trial Registration Data Set | 2 |
| Protocol version | 2 | Version date and identifier | 2 |
| Roles and responsibilities | 3a | Names, affiliations, and roles of protocol contributors | 1 |
| | 3b | Name and contact information for the trial sponsor | 1 |
| | 3c | Role of trial sponsor and funders in design, conduct, analysis, and reporting of trial; including any authority over these activities | 1 |
| | 3d | Composition, roles, and responsibilities of the coordinating site, steering committee, endpoint adjudication committee, data management team, and other individuals or groups overseeing the trial, if applicable | 1 |
| **Open science** | | | |
| Trial registration | 4 | Name of trial registry, identifying number (with URL), and date of registration. If not yet registered, name of intended registry | 2 |
| Protocol and statistical analysis plan | 5 | Where the trial protocol and statistical analysis plan can be accessed | 2 |
| Data sharing | 6 | Where and how the individual de-identified participant data (including data dictionary), statistical code, and any other materials will be accessible | na |

| | | | |
|---|---|---|---|
| Funding and conflicts of interest | 7a | Sources of funding and other support (e.g., supply of drugs) | 14 |
| | 7b | Financial and other conflicts of interest for principal investigators and steering committee members | 14 |
| Dissemination policy | 8 | Plans to communicate trial results to participants, healthcare professionals, the public, and other relevant groups (e.g., reporting in trial registry, plain language summary, publication) | 13-14 |
| **Introduction** | | | |
| Background and rationale | 9a | Scientific background and rationale, including summary of relevant studies (published and unpublished) examining benefits and harms for each intervention | 3-4 |
| | 9b | Explanation for choice of comparator | 6 |
| Objectives | 10 | Specific objectives related to benefits and harms | 4 |
| **Methods: Patient and public involvement, trial design** | | | |
| Patient and public involvement | 11 | Details of, or plans for, patient or public involvement in the design, conduct, and reporting of the trial | 5 |
| Trial design | 12 | Description of trial design including type of trial (e.g., parallel group, crossover), allocation ratio, and framework (e.g., superiority, equivalence, non-inferiority, exploratory) | 5 |
| **Methods: Participants, interventions, and outcomes** | | | |
| Trial setting | 13 | Settings (e.g., community, hospital) and locations (e.g., countries, sites) where the trial will be conducted | 6 |
| Eligibility criteria | 14a | Eligibility criteria for participants | 6 |
| | 14b | If applicable, eligibility criteria for sites and for individuals who will deliver the interventions (e.g., surgeons, physiotherapists) | 6 |
| Intervention and comparator | 15a | Intervention and comparator with sufficient details to allow replication including how, when, and by whom they will be administered. If relevant, where additional materials describing the intervention and comparator (e.g., intervention manual) can be accessed | 6-9 |

| | | | |
|---|---|---|---|
| | 15b | Criteria for discontinuing or modifying allocated intervention/comparator for a trial participant (e.g., drug dose change in response to harms, participant request, or improving/worsening disease) | 13 |
| | 15c | Strategies to improve adherence to intervention/comparator protocols, if applicable, and any procedures for monitoring adherence (e.g., drug tablet return, sessions attended) | 6-9 |
| | 15d | Concomitant care that is permitted or prohibited during the trial | 6-9 |
| Outcomes | 16 | Primary and secondary outcomes, including the specific measurement variable (e.g., systolic blood pressure), analysis metric (e.g., change from baseline, final value, time to event), method of aggregation (e.g., median, proportion), and time point for each outcome | 9 |
| Harms | 17 | How harms are defined and will be assessed (e.g., systematically, non-systematically) | na |
| Participant timeline | 18 | Time schedule of enrollment, interventions (including any run-ins and washouts), assessments, and visits for participants. A schematic diagram is highly recommended (see Figure) | 5 |
| Sample size | 19 | How sample size was determined, including all assumptions supporting the sample size calculation | 9-10 |
| Recruitment | 20 | Strategies for achieving adequate participant enrollment to reach target sample size | 9-10 |
| **Methods: Assignment of interventions** | | | |
| Randomization: | | | |
| Sequence generation | 21a | Who will generate the random allocation sequence and the method used | 10 |
| | 21b | Type of randomization (simple or restricted) and details of any factors for stratification. To reduce predictability of a random sequence, other details of any planned restriction (e.g., blocking) should be provided in a separate document that is unavailable to those who enroll participants or assign interventions | 10 |
| Allocation concealment mechanism | 22 | Mechanism used to implement the random allocation sequence (e.g., central computer/telephone; sequentially numbered, opaque, sealed containers), describing any steps to conceal the sequence until interventions are assigned | 10 |

| | | | |
|---|---|---|---|
| Implementation | 23 | Whether the personnel who will enroll and those who will assign participants to the interventions will have access to the random allocation sequence | 10 |
| Blinding | 24a | Who will be blinded after assignment to interventions (e.g., participants, care providers, outcome assessors, data analysts) | 10 |
| | 24b | If blinded, how blinding will be achieved and description of the similarity of interventions | na |
| | 24c | If blinded, circumstances under which unblinding is permissible, and procedure for revealing a participant's allocated intervention during the trial | na |
| **Methods: Data collection, management, and analysis** | | | |
| Data collection methods | 25a | Plans for assessment and collection of trial data, including any related processes to promote data quality (e.g., duplicate measurements, training of assessors) and a description of trial instruments (e.g., questionnaires, laboratory tests) along with their reliability and validity, if known. Reference to where data collection forms can be accessed, if not in the protocol | 10-12 |
| | 25b | Plans to promote participant retention and complete follow-up, including list of any outcome data to be collected for participants who discontinue or deviate from intervention protocols | 10-12 |
| Data management | 26 | Plans for data entry, coding, security, and storage, including any related processes to promote data quality (e.g., double data entry; range checks for data values). Reference to where details of data management procedures can be accessed, if not in the protocol | 10-12 |
| Statistical methods | 27a | Statistical methods used to compare groups for primary and secondary outcomes, including harms | 12-13 |
| | 27b | Definition of who will be included in each analysis (e.g., all randomized participants), and in which group | 12-13 |
| | 27c | How missing data will be handled in the analysis | 12-13 |
| | 27d | Methods for any additional analyses (e.g., subgroup and sensitivity analyses) | 12-13 |
| **Methods: Monitoring** | | | |

| | | | |
|---|---|---|---|
| Data monitoring committee | 28a | Composition of data monitoring committee (DMC); summary of its role and reporting structure; statement of whether it is independent from the sponsor and funder; conflicts of interest and reference to where further details about its charter can be found, if not in the protocol. Alternatively, an explanation of why a DMC is not needed | 13 |
| | 28b | Explanation of any interim analyses and stopping guidelines, including who will have access to these interim results and make the final decision to terminate the trial | na |
| Trial monitoring | 29 | Frequency and procedures for monitoring trial conduct. If there is no monitoring, give explanation | 13 |
| **Ethics** | | | |
| Research ethics approval | 30 | Plans for seeking research ethics committee/institutional review board approval | 13 |
| Protocol amendments | 31 | Plans for communicating important protocol modifications to relevant parties | 13-14 |
| Consent or assent | 32a | Who will obtain informed consent or assent from potential trial participants or authorized proxies, and how | 13-14 |
| | 32b | Additional consent provisions for collection and use of participant data and biological specimens in ancillary studies, if applicable | na |
| Confidentiality | 33 | How personal information about potential and enrolled participants will be collected, shared, and maintained in order to protect confidentiality before, during, and after the trial | 10-12 |
| Ancillary and post-trial care | 34 | Provisions, if any, for ancillary and post-trial care, and for compensation to those who suffer harm from trial participation | na |

*We strongly recommend reading this checklist in conjunction with the SPIRIT 2025 Explanation and Elaboration and the SPIRIT 2025 Expanded Checklist for important clarifications on all the items. We also recommend reading relevant SPIRIT extensions. See www.consort-spirit.org

Citation: Chan A-W, Boutron I, Hopewell S, Moher D, Schulz KF, et al. SPIRIT 2025 statement: updated guideline for protocols of randomised trials. BMJ 2025;389:e081477. https://dx.doi.org/10.1136/bmj-2024-081477

© 2025 Chan A-W et al. This is an Open Access article distributed under the terms of the Creative Commons Attribution License (https://creativecommons.org/licenses/by/4.0/), which permits unrestricted use, distribution, and reproduction in any medium, provided the original work is properly cited.

#### Supplementary file 2: Social norm feedback intervention design mapped against design recommendations

| GUIDELINES | | APPLICATION TO OUR INTERVENTION DESIGN |
|---|---|---|
| **Focus of intervention** | Care areas that are a priority for the organisation and for patients and are perceived as important by the recipients of the feedback | Antibiotic use was identified as a key quality and safety concern by project partners. Aged care providers have a responsibility to ensure appropriate antibiotic use as per Standards and Infection Prevention and Control Lead roles in RAC. Residents and family/carers want high quality care for residents. Limitation: RAC homes have a lot of competing priorities. |
| | Care areas with high volumes, high risks (for patients or providers), or high costs | There is a high volume of antibiotic use in RAC: 70% of residents will be prescribed a systemic antibiotic annually. Our data show that in any quarter between ~28% and 54% of residents will be prescribed an antibiotic among the project partner homes. Additionally, antibiotic use carries a risk of adverse effects, particularly in the RAC population. |
| | Care areas where there is variation across healthcare providers/organizations in performance and where there is substantial room for improvement | Range of DOT/1000 days in Q4 of 2024 was 17.0 to 160.3. Similarly, for Q1 of 2024, it was 29.8 to 128.1. Antibiotic use is high in RAC and there are high rates of inappropriate use. Only 28.5% of antibiotic courses meet infections requiring antibiotic treatment criteria. Our data also show that course duration is an issue e.g. 91% of trimethoprim courses for women were >3 days. |
| | Care areas where performance on specific measures can be improved by providers because they are capable and responsible for improvements (for example, changing specific prescribing practices rather than changing the overall management of complex conditions) | Prior studies have shown that RAC staff (e.g. registered nurses) play a critical role in communicating with doctors about management of infections. Recommendations will include those to reduce antibiotic initiation, as well as to reduce antibiotic treatment once it is initiated. Decreases in either one of these will reduce the outcome of DOT/1000days. |

| | | |
|---|---|---|
| | Care areas where clear high-quality evidence about best practice is available | Therapeutic Guidelines contain clear recommendations about antibiotic treatment including course length and when treatment is not warranted. A limitation in the RAC population is that diagnosing infections in some residents is complicated by other comorbidities, including dementia, and there is substantial 'just in case' treatment. |
| **Audit component** | Indicators include relevant measures for the recipient (this may include structure, processes and/or outcomes of care, including patient-reported outcomes) that are specific for the individual recipient | Two measures will be presented: days of therapy (DOT)/1000 resident days and the percentage of courses longer than guidelines. Feedback will be at home level (residential manager) which is the key unit of RAC organisation. |
| | Indicators are based on clear high-quality evidence (for example, guidelines) about what constitutes good performance | DOT/1000 days is an accepted measure of antibiotic use. Guidelines on antibiotic treatment will be presented as part of recommendations. |
| | Data are valid and perceived as credible by the report recipients | Data are extracted from medication administration systems of RAC providers and a reflection of medication residents are administered. Supporting information is drawn from nationally recognised treatment guidelines and referenced. DOT/1000 days is case mix adjusted to support fair comparisons between RAC homes. |
| | Data are based on recent performance | Feedback will be sent quarterly and present data for the last quarter with a maximum 2-week lag. |
| | Data are about the individual/team’s own behaviour(s) | Data will be about each RAC home’s performance. |
| | Audit cycles are repeated at a frequency informed by the number of new patient cases with the condition of interest such that new audits can capture attempted changes | Feedback to be provided quarterly. Our data show that in any quarter between ~28% and 54% of residents will be prescribed an antibiotic among the project partner homes |
| **Feedback component** | Presentation is multimodal including either text and talking or text and graphical materials | Data will be presented in text and graphically. The DOT/1000 days measure will be explained in text and graphically in supporting information. |

| | | |
|---|---|---|
| | Delivery comes from a trusted, credible source (for example, supervisor or respected colleague), with open acknowledgement of potential limitations in the data | Signatory of the feedback to be respected leader with each RAC provider. |
| | Feedback includes a relevant comparator to allow the recipient to immediately identify if they are meeting the desired performance level | Comparison rate will be provided for DOT/1000 days and will comprise of top-performing peers, defined as the mean of the lowest 10% of the RAC homes. |
| | A short, actionable declarative statement should describe the discrepancy between actual and desired performance, followed by detailed information for those interested | Brief text on discrepancy will be provided. Comparison group will be labelled as 'Best performers'. Message will be varied depending on each home's performance relevant to the target. |
| **Targets, goals and action plan** | The target performance is provided; the target may be based on peer data or on a consensus-approved benchmark | Target performance DOT/1000 days will be provided consisting of the top performing peers in corresponding feedback period. |
| | Goals for target behaviour are specific, measurable, achievable, relevant and time-bound | The feedback contains clear explanations on how RAC homes can improve their performance. Measures have been generated on a quarterly basis for the aged care provider leadership teams and the feedback reports are designed for the RAC home teams. |
| | A clear action plan is provided when discrepancies are evident | Feedback will be supported with clear actions to reduce antibiotic initiation and reduce treatment length, with links to resources and guidelines. The percentage of courses longer than guidelines will in the last quarter will be presented to support actions. |
| **Organizational context** | Audit and feedback is part of a structured programme with a local lead | Each RAC provider has been involved in co-developing the program and have provided signatories from within the organisation's leadership team. |
| | Audit and feedback is part of an organizational commitment to a constructive, non-punitive approach to continuous quality improvement | Feedback includes message that the program is part of quality improvement and that the aged care provider is committed to improving antibiotic use. |

| | | |
|---|---|---|
| | Recipients have or are provided with the time, skills and/or resources required to analyse and interpret the data available | Explanations of the data presented are included in the feedback report to ensure data and suggested actions are understood by recipients. Trusted guidelines and government resources are also linked and included. |
| | Teams are provided with the opportunity to discuss the data and share best practices | It is envisaged that feedback will be discussed at Medication Advisory Committee meetings. |

**Supplementary file 3: Semi-structured interview questions for RAC staff and consumers.**

**Audit and feedback (these questions for RAC staff):**

1. Did you receive a letter/email with feedback about antibiotic use in your home? If Yes:
    - Did you open the letter/email, read the information?
    - Was it easy to understand?

2. What was your overall impression of the feedback you received?
    - Which parts of the feedback were most/least useful or informative?
    - Do you think the information about antibiotic use in your home was accurate? Why/why not?
    - Was the data included in the letter (i.e. data from the previous quarter) still relevant to you?

3. How did you use the information presented in the feedback?
    - Seek additional information from internal/external sources
    - Change or reconsider actions, e.g. dipstick testing; requesting antibiotics for bronchitis, viral respiratory infections; review duration of use
    - If No: why not?

4. Did you share the feedback with others?
    - e.g. MAC, facility staff, pharmacist, GPs, residents?

**Public commitment poster (these questions for RAC staff, consumers - have a copy of poster to show participant)**

1. Have you seen this poster displayed in your home? If yes, use prompts:
    - Where/when did you see it?
    - What did you think of the design, visuals?
    - Was it in a location where stakeholders (i.e. staff, residents, residents' families) would be likely to see it?

2. Was the information in the poster easy to understand?
    - Was there any information that was new/unfamiliar to you?

3. Have you discussed the poster with anyone? Prompt: staff, pharmacist, GPs, residents?
    - If Yes: Who was it with? Was the discussion useful?
    - If No: Why not?

4. Did you take any other action after looking at the poster?
    - Prompts for staff: seek additional information about antibiotic use, change practices e.g.: dipstick testing, review duration of use.
    - Prompts for consumers: seek additional information about antibiotic use, query prescriptions and/or duration of use when receiving antibiotics.
    - If No: Why not? What would make it more likely to take action?

**Consumer information (these questions for consumers (residents and families – have a copy of resources to show participant):**

1. Have you seen this leaflet/flyer/information displayed in your home? If yes, use prompts:
    - Where/when did you see it?
    - What did you think of the design, visuals?
    - Who do you think the target audience is?

2. Were the messages easy to understand?
    - Was there any information that was new/unfamiliar to you?

3. Have you discussed the information with anyone? Prompt: family member, residents, staff, pharmacist, GP?
    - If Yes: Who was it with? Was the discussion useful?
    - If No: Why not??

4. Did you take any other action after seeing this information?
    - If Yes: Prompts: seek additional information about antibiotic use, query prescriptions and/or duration of use when receiving antibiotics.
